# Medical Expectations Survey on Artificial Intelligence Solutions in daily practice

**DOI:** 10.1101/2023.06.29.23291561

**Authors:** Mara Giavina-Bianchi, Edson Amaro, Birajara Soares Machado

**Author notes:** **CORRESPONDING AUTHOR:** Mara Giavina-Bianchi.

## Abstract

**Background:** Artificial intelligence (AI) applied to Medicine has become one of the hottest topics for the past years. Although scarcely used in real practice, it brings along many expectations, doubts and fears for physicians. Surveys can help to understand this situation.

**Objective:** To explore the degree of knowledge, expectations, fears and daily practice questions on AI use by physicians.

**Methods:** an electronic survey was sent to physicians of a large hospital in Brazil, from August-September 2022.

**Results:** 171 physicians responded to our survey. 54% considered themselves to have an intermediate knowledge of AI. 79% believe AI should be regulated by a Governmental Agency. If AI were reliable and available, 78% intend to use AI frequently/always for diagnosis (87%) and/or management (83%), but they were unsure about the use of AI by other health professionals (50%) or by the patients (51%). The main benefit would be increasing the speed for diagnosis and management (64%), and the worst issue, to over rely on AI and lose medical skills (71%). Physicians believe AI would be useful (94%), facilitate the work (87%), increase the number of appointments (54%), not interfere in the financial gain (58%) and not replace their jobs, but, rather, be utilized as an additional source of information (65%). In case of disagreement between AI and physicians, most answered that a third opinion should be requested (86%). There were no significant differences between the physicians’ answers according to time since graduation.

**Conclusions:** physicians showed to have good expectations regarding the use of AI in Medicine when applied by themselves, but not so much by others. They also have intention to use it, as long as it was approved by a Regulatory Agency. Although there was hope for the beneficial impact of AI on healthcare, it also brings specific concerns.

## 1) INTRODUCTION

The use of artificial intelligence (AI) is expanding throughout the field of Medicine, driven by researchers and entrepreneurs(1, 2). Over the last decade, the number of publications on AI in medicine and biomedicine have substantially increased(3). AI solutions might change the clinical practice in virtually all medical disciplines and areas of health care. Despite the potential of machine learning to improve multiple aspects of patient care, there are still barriers to clinical adoption. Important questions remain regarding how machine learning interventions are being incorporated into health care(4). A reluctance to adopt AI-based solutions might be due to a lack of knowledge, fear of error and concerns about losing jobs and/or power(5). Some other perceived limitation of AI applications is the belief that communication and empathy are human competencies that cannot be replaced by AI. Also, the ability to provide value-based care needs the physicians’ judgments. Some possible benefits included expectations about improved efficiencies, specially, with respect to the reduction of administrative burdens on physicians(6). Examples of practical use of AI solutions in clinical routine are still scarce around the globe(2). The increasing development in medical systems using AI brings enormous expectations and fears for both physicians and patients.

Physicians are likely to be the “earliest” adopters of AI solutions for patient care and inevitably should become direct AI operators. Therefore, they play a pivotal role in the acceptance and implementation of clinical AI, and consequently their views need to be known, explored, and understood(7). Opinion surveys are important tools in assessing satisfaction with a particular service and consist of a list of questions whose objective is to extract certain data from a group of people(8). Previous studies on the acceptance of the use of AI in Medicine were limited to specific areas, such as radiology(5, 9-11), dermatology(1, 12-14), ophthalmology (15-17), and, also, to specific countries. However, at the time of this writing, we were not able to find studies exploring this subject in Brazilian physicians, and very few in Latin America – leaving a gap in this part of the globe: is the perception of AI adoption similar to other countries? And at the same time, one aspect yet not explored, does the acceptance of AI solutions vary according to the number of years since medical graduation? Routinely observation indicates that younger individuals are keener to accept new technologies.

The main objective of this study was to assess the expectations, fears, and thoughts of Brazilian physicians about some practical aspects of the hypothetical use of AI solutions in medical daily practice. The secondary objective was to verify if there were differences in the opinions between physicians according to the time since their graduation.

## 2) METHODS

We performed a cross-sectional observational study via an opinion survey study approved by the Ethics Committee of Hospital Israelita Albert Einstein (CAAE: 30749620.6.0000.0071). The surveys were sent to 7,457 physicians from the clinical staff linked to Hospital Israelita Albert Einstein (HIAE) and there were no exclusion criteria and responses collected from 08/01 to 10/01/2022 (60 days). HIAE is a non-profit, public interest organization based in São Paulo, Brazil. There was no financial incentive to answer the questionnaire.

### 2.1 Questionnaire

The questionnaire was composed by 30 questions. Question number one was the Informed Consent Form (ICF). In case of acceptance of the ICF, the next questions were presented to the individual. If not, the survey would be terminated. The time to complete the questionnaire was approximately 7 minutes on average. The survey was completely anonymous and confidential, and only the authors of the work had access to the answers. The complete questionnaire can be viewed in Table 1SM (Supplementary Material). The questions were developed by the authors based on their experience in developing AI solutions for physician and on previous published medical literature. It was divided into six sections. The first one was the Informed Consent Term (question 1). Sections 2 (questions 2-12) and 3 (questions 18-20) were aimed to investigate physicians’ profile and possible utility of IA medical solutions, respectively. Sections 4 (questions 21-24) and 5 (questions 25-30) were related to expected benefits, problems, financial issues, possible disagreements, and legal aspects. Along with the questions, there were many opportunities for physicians to make comments in an open box about the answers.

**Table 1:**
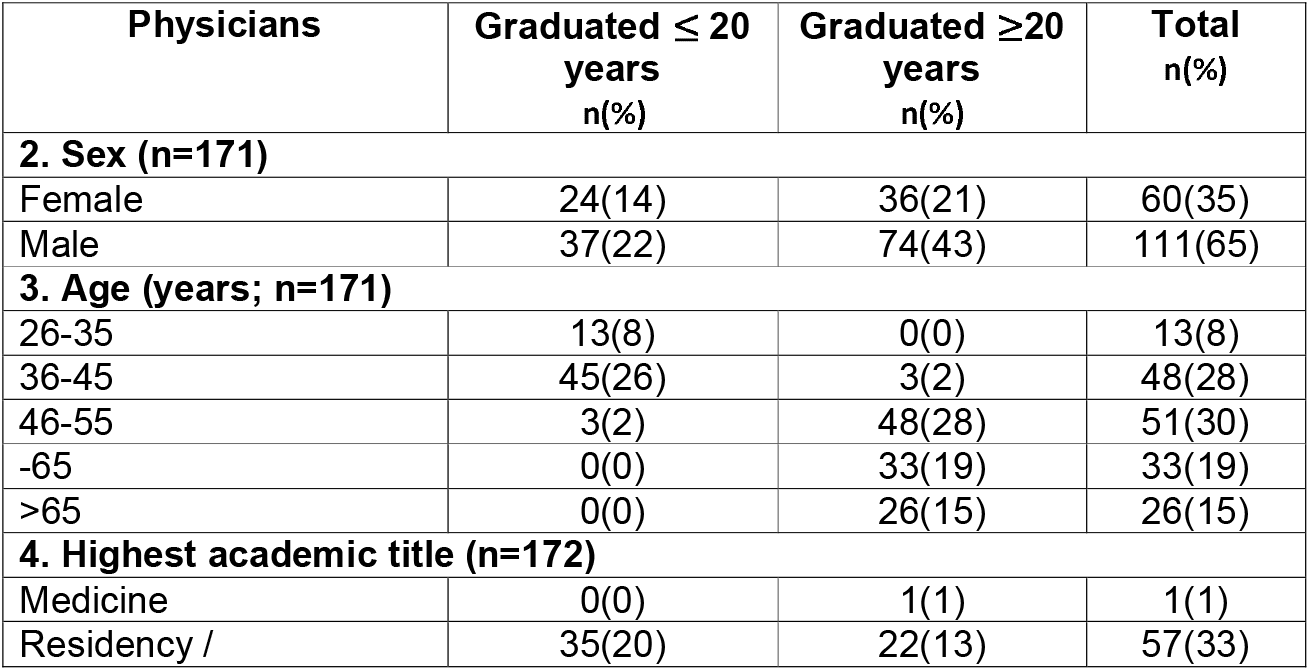
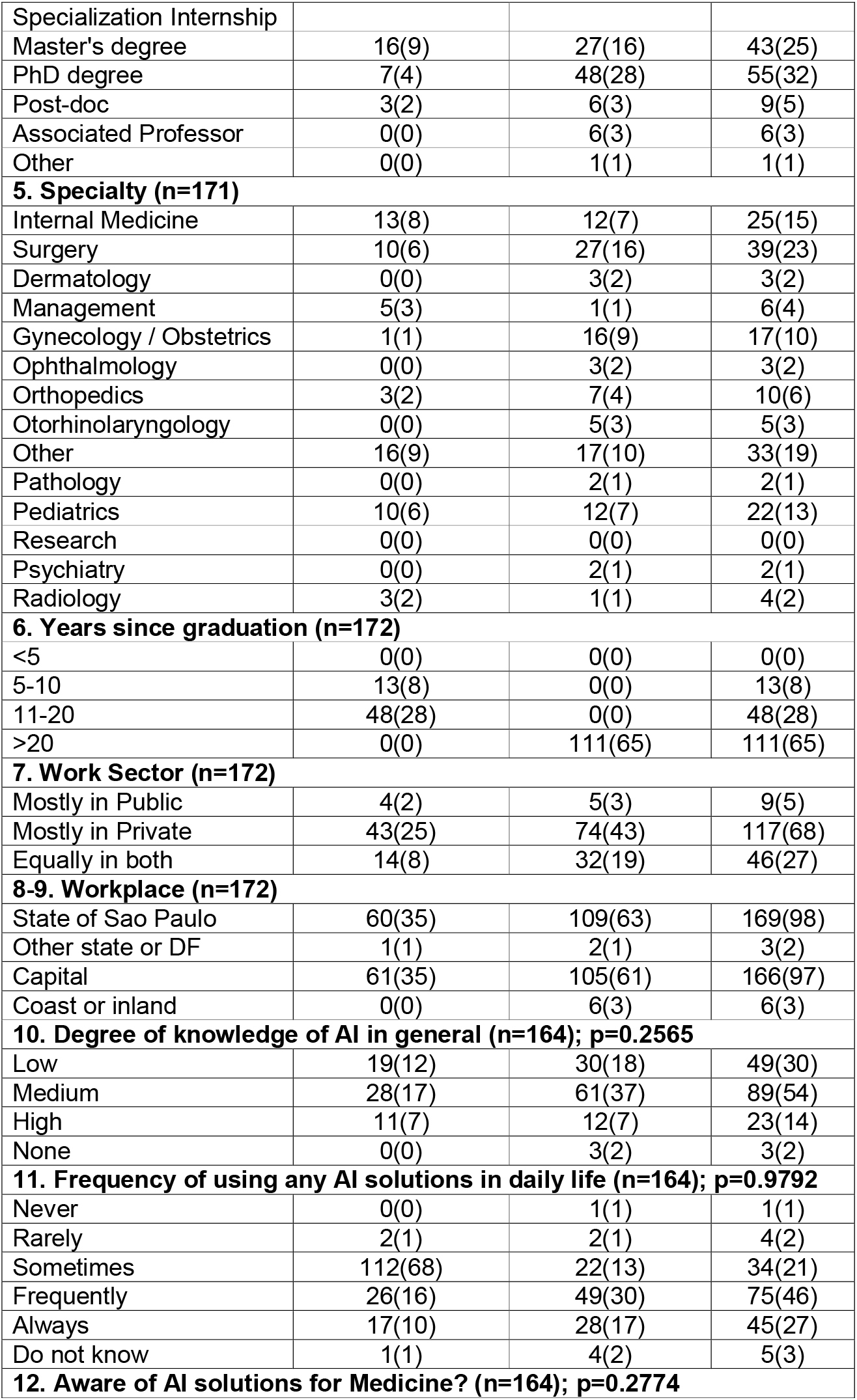
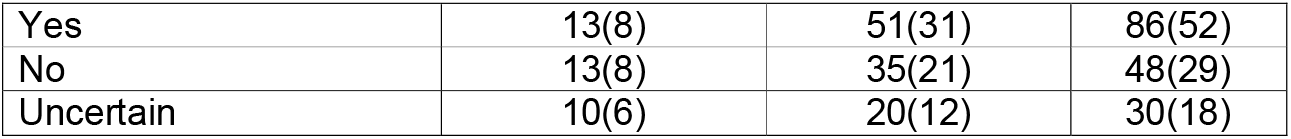
Profile of physicians who answered the opinion questionnaire on artificial intelligence solutions at HIAE (answers for questions 2-12).

### 2.2 E-Mailing the questionnaire

The survey was sent by e-mail to all physicians with an electronic address linked to the HIAE. In the first email, on 08/01/2022, a brief introduction inviting the physician to participate in the survey and the questionnaire link to be completed in the *SurveyMonkey computer program* (SurveyMonkey Inc., San Mateo, CA, USA; www.surveymonkey.com) were sent to all physicians. In the second round (08/08/2022), we replicated the same email to those who had not visualized the previous one. There was a third email sent on 09/05/2022, to the remaining individuals. The survey ended on 09/30/2022. This time framing of 60 days and the number of reminders during the period followed the current *modus operandi* of the hospitals’ marketing department for all studies sent electronically. *SurveyMonkey’s* program has a blocking mechanism that prevents the same subject to respond to the survey more than once. It identifies and notifies the user that the questionnaire had already been answered, blocking a new response. The research was previously tested on 3 physicians of the HIAE medical team, who were part of the tested target population. Our work followed the guide to reporting CROSS survey studies (Checklist for Reporting Survey Studies)(4).

### 2.3 Statistical analysis

Response rate was calculated by the number of physicians who responded the questionnaire divided by the number of physicians to whom the e-mail was sent to x 100. Completion rate was the number of surveys answered and sent/number of surveys initiated by respondents x 100. The subjects were divided into 2 groups: medical school graduation for ≤20 years or >20 years, according to the answer to question 7 of the questionnaire. Statistical analyses between the 2 groups were performed using the chi-square test in Prism software version 6 (GraphPad Software, Inc., San Diego, CA, USA). P value <0.05 was considered significant.

## 3. RESULTS

### 3.1 Descriptive results

Acceptance of the ICF was performed by 181 physicians. The response rate was 2.4 % (181/7457). The completion rate of the questionnaire was 91% (164/181). Table 1 shows the profile of the physicians who responded to the survey. They were mostly men, from the private sector, 46-55 years of age and 20 or more years since medical graduation. As for the place of work, 117 (68%) said they work mainly in the private sector; 46 (27%), equally in both sectors, and 9 (5%), mainly in the public. We see that all academic degrees were present in the study (the majority with residency/specialization, PhD or Master). The distribution among specialties were heterogeneous and skewed, but it may reflect the different number of physicians of each specialty linked to the hospital mailing list. There are probably many more pediatricians in the hospital’s mail list than psychiatrists. Almost the entirety of them works in São Paulo city, as the main hospital is in this location. Most of them classified their knowledge of AI as intermediate. They frequently use AI algorithms in general and claim to be aware of AI algorithms applied to Medicine.

### 3.2 Main results

In total 164 subjects were consistent in answering the questionnaire until the end. Figure 1 presents answers from questions 13-17 of the survey. They believe AI would be helpful for patient’s diagnosis, management and to support image exams interpretation. They are not so certain about the use of AI by other health professionals, such as nurses or physiotherapists, or by the patient themselves.

**Fig. 1:**
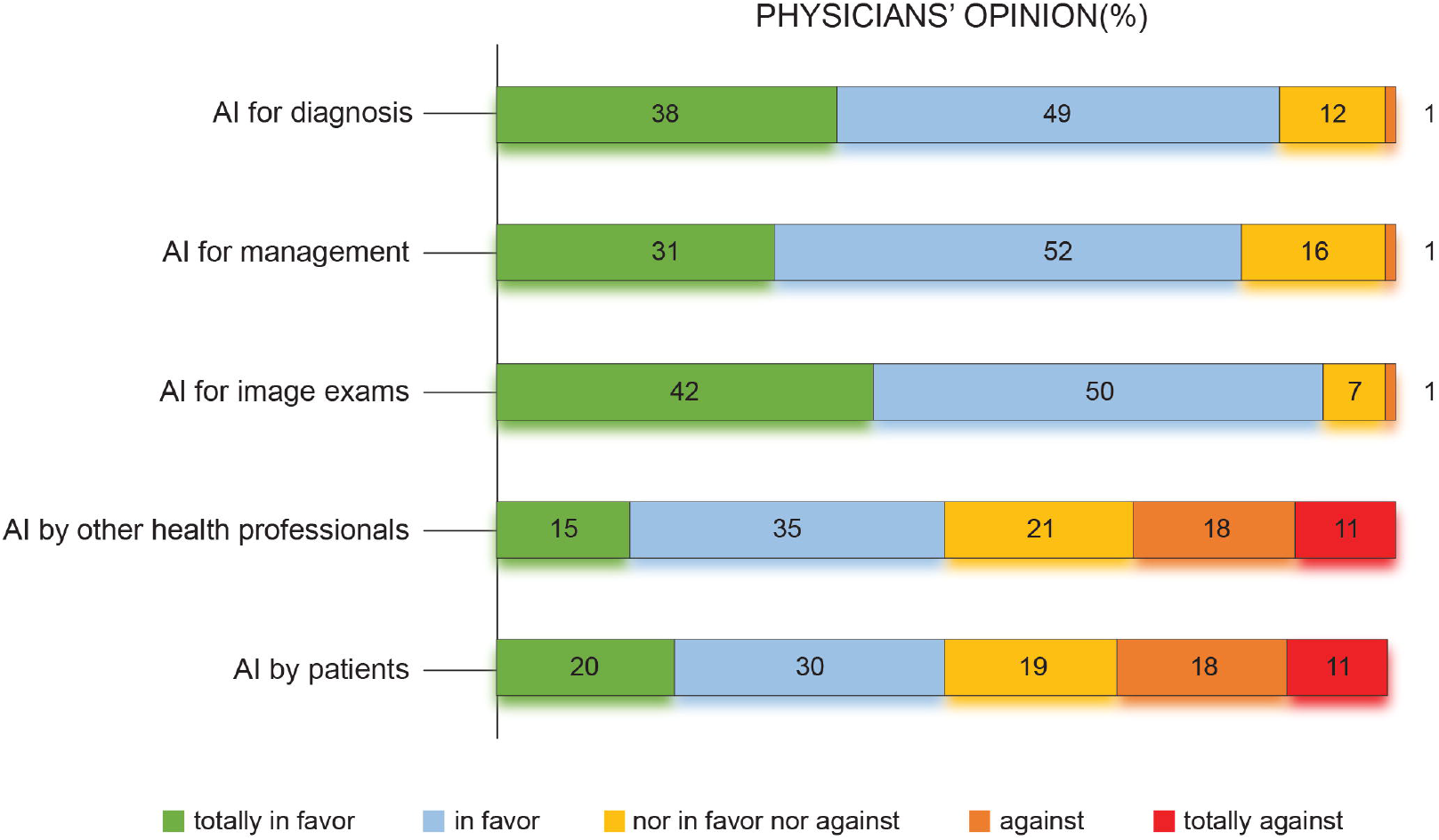
Physicians’ expectations about the role of artificial intelligence solutions used by themselves or by other health care professionals or patients (questions 13-18).

If filling the patient’s shoes, physicians acknowledge that AI diagnosis solutions used by non-specialists might cause distress about some types of diagnosis, such as skin melanoma (question 18). This question asked if a melanoma detection solution was used by the physicians on themselves in a certain lesion, and the AI showed a high probability of melanoma diagnosis; what that situation would elicit in their feelings (the degree of anxiety or none) and what action would be taken (how quick would like to seek for a specialist appointment or not). The major benefits cited by the physicians were greater speed for diagnosis and management, greater accuracy, healthcare cost reduction and greater access. The main issues listed were the fear to rely excessively on the AI algorithms driving the physicians to lose their medical skills, wrongful diagnostic or management reports and increasing the distance in the medical-patient relationship (Table 2).

**Table 2:** Overview and expectations about artificial intelligence solutions in general and in Medicine of medical work among those who answered the opinion questionnaire at HIAE (questions 18-20)

| Questions 18-20 | ≤20 years<br>n(%) | >20 years<br>n(%) | Total<br>n(%) |
| --- | --- | --- | --- |
| **18. Consequences of AI diagnosis as a patient (p.e. suspicious melanoma) (n=160); p=0.3792** |  |  |  |
| Extremely anxious/ immediate appointment | 36(23) | 64(40) | 100(63) |
| Anxious/ appointment whenever possible | 15(9) | 28(18) | 43(27) |
| Not shaken/ appointment whenever possible | 4(3) | 9(6) | 13(8) |
| Not shaken/no appointment | 1(1) | 0(0) | 1(1) |
| I am dermatologist | 0(0) | 3(2) | 3(2) |
| **19. AI expected benefits (n=162; total choices = 437); p=0.7272\*** |  |  |  |
| Greater speed | 42(10) | 64(15) | 104(24) |
| Greater accuracy | 33(8) | 54(12) | 85(19) |
| Cost reduction | 29(7) | 58(13) | 85(19) |
| Reduction in the number of subsidiary exams | 21(5) | 34(8) | 53(12) |
| Reduction in patient's anxiety | 1(0) | 8(2) | 9(2) |
| Greater access to healthcare | 27(6) | 42(10) | 67(15) |
| Greater patient's participation in healthcare | 9(2) | 22(5) | 29(7) |
| Other | 2(0) | 3(1) | 5(1) |
| **20. AI expected problems (n=163; total choices= 462); p=0.2138\*** |  |  |  |
| Confidentiality issues | 11(2) | 20(4) | 31(7) |
| Worsening medical-patient relationship | 27(6) | 59(13) | 86(19) |
| Wrongful use of patient's information by employers and health companies | 21(5) | 47(10) | 68(15) |
| Errors in diagnosis/ management | 31(7) | 55(12) | 86(19) |
| Physicians relay too much on AI and lose medical skills | 44(10) | 74(16) | 118(26) |
| Healthcare increase cost | 9(2) | 10(2) | 19(4) |
| Lack of transparency | 17(4) | 28(6) | 45(10) |
| Other | 7(2) | 2(0) | 9(2) |

\*physicians could choose more than one benefit and problem, thus, the % was calculated taking into account the total number of choices made in each question.

Overall, physicians intend to apply AI in Medicine frequently, they believe it will facilitate their work, will not interfere with the number of appointments and it would be useful for patient’s diagnosis and management (Figure 2).

**Fig. 2:**
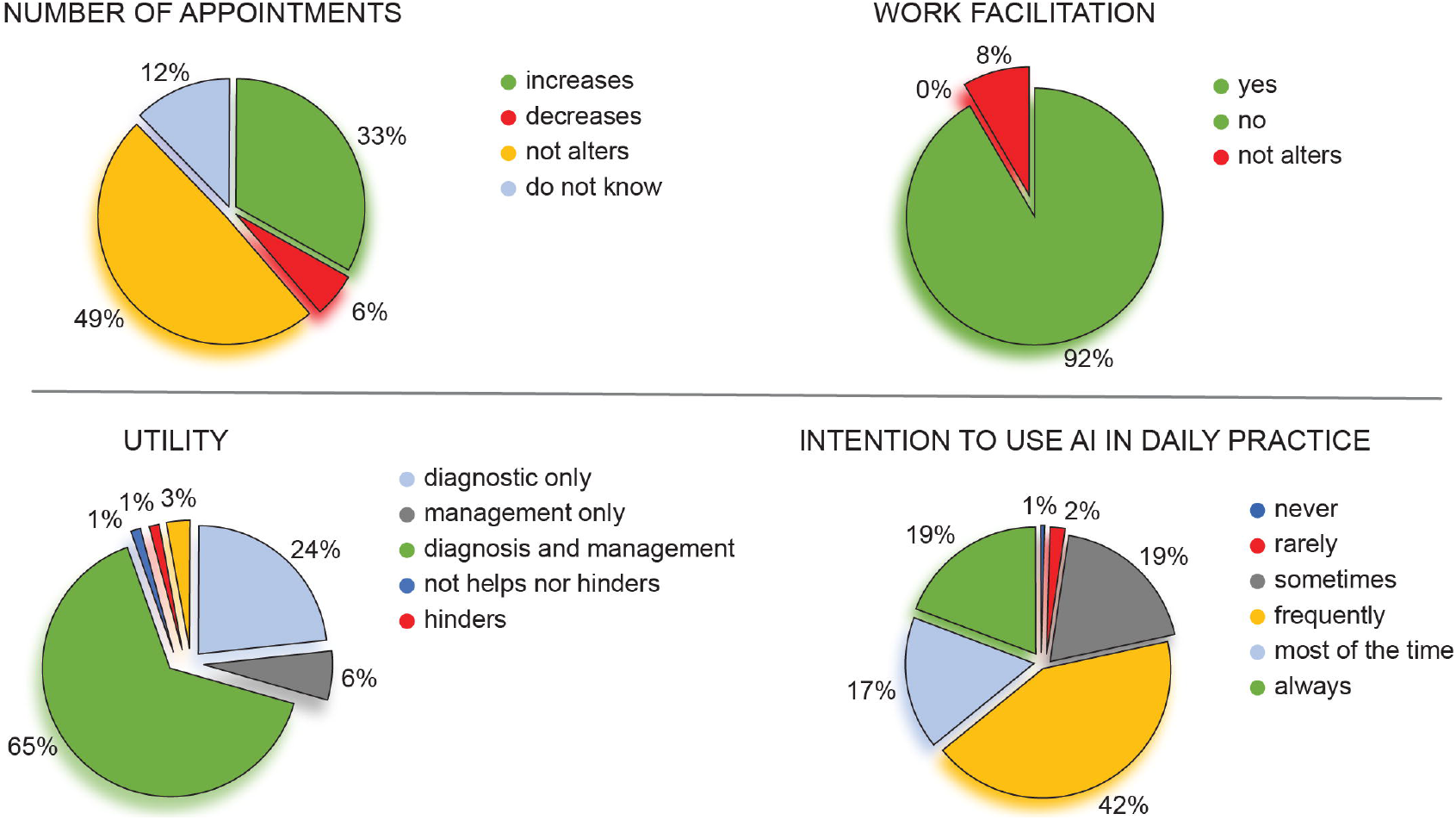
Questions 21-24 showing the intention of use artificial intelligence solutions in daily practice, idea of work facilitation, interference with number of appointments and possible utility proposals.

Table 3 details the main answers for questions 25-30, assessing AI algorithms will not substitute the physician, but be one more source of information and will not alter their financial gain. In the event of a diagnosis or conduct disagreement between the physicians and the AI solution, we proposed 2 scenarios. In the first one, AI algorithms and physicians had the same accuracy rate for a defined task, and, in the second, AI had usually a better performance rate than physicians. What should be done in each case? For the former case, they were a bit divided between “asking for a third opinion” or “the medical opinion should be followed”. In the latter situation, the great majority chose to request a third opinion. As for legal responsibility, most individuals answered that it should be shared between AI algorithm’s manufacture and physicians/hospitals. 79% responded that AI solutions should have the stamp of a regulatory Governmental Agency. Finally, no statistical differences in answers were found between physicians who graduated for ≤20 years and >20 years.

**Table 3:** Effects of artificial intelligence solutions on the routine of medical work among those who answered the opinion questionnaire at HIAE (questions 25-30)

| Questions 25-30 | $\leq 20$ years<br>n(%) | $>20$ years<br>n(%) | Total<br>n(%) |
| --- | --- | --- | --- |
| **25. Physician Replacement in Specialties Image based? (n=163); p=0.4050** |  |  |  |
| Totally | 3(2) | 2(1) | 5(3) |
| Partially | 17(10) | 34(21) | 51(31) |
| Be one more source | 37(23) | 68(42) | 105(64) |
| Not alter | 1(1) | 0(0) | 1(1) |
| Do not know | 0(0) | 1(1) | 1(1) |
| **26. Financial gain? (n=163); p=0.5500** |  |  |  |
| Increases | 10(6) | 12(7) | 22(13) |
| Decreases | 8(5) | 18(11) | 26(16) |
| Not alter | 33(20) | 61(37) | 94(58) |
| Do not know | 7(4) | 14(9) | 21(13) |
| **27. Disagreement between AI and physicians (equal accuracy rates)? (n=162); p=0.4644** |  |  |  |
| Should favor physician's | 28(17) | 44(27) | 72(44) |
| Should favor IA's | 0(0) | 1(1) | 1(1) |
| Request third opinion | 30(19) | 56(35) | 86(53) |
| Do not know | 0(0) | 3(2) | 3(2) |
| **28. Disagreement between AI and physicians (AI greater accuracy rate)? (n=160); p=0.2146** |  |  |  |
| Should favor physician's | 13(8) | 16(10) | 29(18) |
| Should favor IA's | 8(5) | 10(6) | 18(11) |
| Request third opinion | 37(23) | 71(44) | 108(68) |
| Do not know | 0(0) | 5(3) | 5(3) |
| **29. Legal Liability (n=159); p=0.1394** |  |  |  |
| Physician only | 13(8) | 32(20) | 45(28) |
| AI only | 2(1) | 6(4) | 8(5) |
| Shared equally | 39(25) | 49(31) | 88(55) |
| Do not know | 4(3) | 14(9) | 18(11) |
| **30. Governmental Regulation? (n=163); p=0.5858** |  |  |  |
| Yes | 43(26) | 85(52) | 128(79) |
| No | 7(4) | 10(6) | 17(10) |
| Do not know | 8(5) | 10(6) | 18(11) |

## 4. DISCUSSION

We conducted a web-based survey study among hospital physicians in a large hospital in Brazil seeking for their opinion in the use of AI solutions in medical practice. To our knowledge, this is the first survey to interrogate physicians’ expectations, fears, and opinions of AI usage in Medicine in Brazil. There was a low rate of response, but according to HIAE marketing department, this was considered a regular respond rate, since physicians usually do not have respond to questionnaires, in general. We wanted to extend the time frame by couple of months for the study, but the department responsible for medical communication is concerned about overwhelming the physicians with too many electronically information. Thus, we followed the hospitals regular modus operandi. The choice of dividing the groups in < or > 20 years since medical graduation was based on our personal experience and the common knowledge that younger individuals are keener to use technology applications than older ones, in general. Thus, individuals > 20 years since medical graduation would be older and might have a different approach towards AI applied to Medicine than younger ones.

Our responders were mostly from the private sector, men, from different medical areas and different ages, which was important to investigate the general perception of medical AI and if physicians more recently graduated were more prone to accept AI solutions than physicians longer graduated. Most of the studies in medical AI acceptance are focused in specific areas, such as radiology(5, 6, 7, 8), dermatology(1, 9, 10, 11) and ophthalmology(12, 13, 14), which could be more affected than others by adoption of AI solutions. Thus, it is important to have more general views about the topic. Our group of responders was very heterogeneous but may reflect the frequency of different specialties in the mailing list of the hospital. Thus, it would be expected to have more answers from pediatricians than from psychiatrists, as example.

Our results demonstrate that, overall, physicians have positive expectations about the use of AI in clinical practice, but also some concerns. Their answers indicate that medical use of AI solutions hopefully will facilitate their work and be useful for diagnoses, treatment, and exams’ interpretation. They believe the number of appointments performed by them, overall, would increase, probably by increase of speed in making diagnosis or managements. Even so, they indicated that AI solutions would not interfere with their financial gain.

Probable benefits of AI solutions included greater speed, accuracy, and cost reduction for the healthcare system. That is in accordance with previous studies. A recent systematic review that included 45 studies with physicians or medical students on clinical AI showed >60% responders with optimistic outlooking in 84% of the studies(3). There is also an expectation that AI in medical practice will meet higher expectations of medical treatment and physicians and will increase the efficiency of clinical care, perceived as the next big thing that will sustainably change medicine towards precision and personalized medicine(15).

Our responders believe they will not be replaced by AI, but it will be one more source of information to support their work. Although the current discourse in medical literature has shifted from replacement to support medical activities, as in the idea of Augmented Intelligence, where humans and AI are together in functions that each of them do best(16), the adoption of AI also opens the possibility of transferring decision making to other health professionals or patients. This probability divides their opinion, with roughly half of them are against it. Many comments revealed the fear of misinterpreting the results if no medical supervision was performed. Question 18 explored the effect of AI used by the physicians in the role of patients themselves, and clearly showed that it can generate a great deal of anxiety if a troublesome diagnosis has been appointed by AI without a specialist supervision. In one article focusing on patients’ opinion about the use of medical AI, they appeared to be receptive to the use of AI for it if implemented in a manner that preserves the integrity of the human physician-patient relationship(1). A review article on the convergence of human and artificial intelligence poses an important statement on that matter: “Over time, marked improvements in accuracy, productivity, and workflow will likely be actualised, but whether that will be used to improve the patient-doctor relationship or facilitate its erosion remains to be seen” (17). In China, another study showed that the general population is more distrustful of AI in medicine unlike the overall optimistic views posed in the AI, and that the level of trust is dependent on what medical area is subject to scrutiny(18). Those aspects are also a big concern for our physicians: worsening of patient-physician relationship was listed right after the fear of over-relying on medical AI, causing them to lose their medical skills over time. Comments about the benefit of human contact and the detection of emotions by the physicians cannot yet be replaced. A study with more than 1,000 physicians showed that the fear of medical AI was inversely associated with advanced or intermediate AI-specific knowledge compared with those with basic(6).

Possible disagreements between AI algorithms and physicians in daily practice were also explored by the questionnaire. In both questions 27 and 28, physicians believed a third opinion should be requested (54% and 70%, respectively). Nevertheless, as the accuracy of the AI outperforms the physician’s in question 28, the number of subjects who answered that the final decision should be the physician’s drops from 45% to only 19%, revealing that the informed performance of AI solutions is crucial for physicians to make decisions.

As for legal aspects, most physicians believe the liability should be shared between them and the AI solution, reflecting the idea that it involves a serious action, which requires careful engagement of all stakeholders. According to a recent article, all players in this field, such as physicians, developers and health care administrators should recognise that the implementation of an AI solution is not just a technical challenge, but rather, presents ethical, legal and social challenges as well. Thus, it is important to gather all stakeholders to develop AI collaboratively from outset to implementation and evaluation(19). It is very clear also that physicians require and trust the role of Government Agencies to regulate this field. This is corroborated by another study, discussing how this will become increasingly important as more algorithms start to be used in real life. A regulatory approval should not only mitigate possible harms, but also define a proper balance between risks and benefits, promote effective validation standards in real settings and innovation(20).

We found no significant differences between the groups of physicians who graduated up to 20 years or > 20 years, which demystifies the popular idea that physicians with more years of practice – likely of older age -would be less prone to adopt new technologies.

In conclusion, our survey explored the physicians view in AI medical solutions in a new global geographical area, showing a general positive attitude towards AI them, but also some concerns, mostly related to agencies regulation and to who they should be using them. No difference between time since graduation groups was seen.

## 5) LIMITATIONS

It is important to notice that our online survey was a cross-sectional study and was based on physicians’ response from one single institution that has a particular interest in innovation and AI in Medicine. Since the study was performed via email, physicians who answered the questionnaire could already be more likely to use technology, in general. Also, the theme of the study was stated in the email subject, so, those interested in it, would be more likely to open the email and answer the questionnaire than those who don’t. Therefore, the results have to be considered within possible bias to a more positive attitude towards technology and AI in healthcare if compared to all physicians working in Brazilian hospitals.

## Data Availability

All data produced in the present study are available upon reasonable request to the authors

## 6) AUTHORS’ CONTRIBUTION

MGB and BM researched literature and conceived the study. MGB, BM and EA were involved in protocol development, gaining ethical approval, patient recruitment and data analysis. MGB wrote the first draft of the manuscript. All authors reviewed and edited the manuscript and approved the final version of the manuscript.

## 7) ACKNOWLEDGEMENTS

many thanks to Isabela Polizel Bonfá, who was responsible for sending the emails to the physicians.

## 9) SUMMARY TABLE

- The use of AI brings many expectations, doubts and fears for physicians
- Physicians showed good expectations for the use of AI if applied by themselves
- They have intention to use it frequently if approved by a Regulatory Agency
- Although hoping for a beneficial impact on healthcare, they have specific concerns

